# EchoGraph System for Automated Quality Assessment of Echocardiography Reports

**DOI:** 10.1101/2025.05.07.25327158

**Authors:** Chieh-Ju Chao, Jean-Benoit Delbrouck, Mohammad Asadi, Imon Banerjee, Juan M. Farina, Francesca Galasso, Ahmed K Mahmoud, Mohammed Tisser Abbas, Yu-Chiang Wang, Reza Arsanjani, Garvan C Kane, Jae K Oh, Bradley J Erickson, Li Fei-Fei, Ehsan Adeli, Curtis Langlotz

**Author notes:** **Address of Correspondence:** 200 1st Street SW, Room: Gonda 4-478, Rochester, MN 55095, USA, Chieh-Ju Chao, MD, Assistant Professor of Medicine, Department of Cardiovascular Medicine, Mayo Clinic, Twitter: @chiehjuchao1.

## Abstract

Generative AI needs automatic clinical text accuracy metrics, but none exist for echocardiography. To address this, we developed EchoGraph, a BERT-based model trained on 600 densely annotated echocardiography reports from the Mayo Clinic (2017), split 7:2:1 for training, validation, and testing, using a tailored schema with 48,256 entities and 29,731 relations annotated. Sixty random MIMIC-EchoNote reports were annotated (3,672 entities and 2,360 relations) for external validation. EchoGraph demonstrated strong performance predicting entities (micro F1 0.85) and relations (micro F1 0.70), maintaining performance on external validation (entity micro F1 0.80, relation micro F1 0.52). EchoGraph F1 score showed superior error sensitivity versus RadGraph F1, with 2.8-fold higher slope magnitude (−0.817 vs −0.291) and better variance explained (R^2^ = 0.803 vs 0.578). EchoGraph offers an effective solution for evaluating language model-based echocardiography applications, supporting more accurate AI-generated reports.

## Introduction

Echocardiography (echo) reports are the primary communication tool for echo studies, containing detailed quantitative measurements and cardiologists’ interpretations concerning each patient’s medical history and clinical context^1,2^. Although these reports hold substantial clinically relevant information, their free-text format presents significant challenges for large-scale use. The unstructured nature of this data, combined with complex medical language, hampers automated extraction and analysis, limiting their utility in artificial intelligence (AI) research and clinical practice^3^. With the rise of generative AI, particularly language models^4,5^, there is an increasing need for automatic, generic metrics to evaluate the factual correctness of generated clinical text to facilitate the pace of progress in developing vision-language models that produce accurate reports^3,6,7^. Studies have leveraged information extraction or language model techniques for the development of automatic metrics or frameworks to evaluate the reliability and accuracy of AI-generated clinical narratives, enhancing their utility in medical practice and research^3,6,8–10^.

However, studies specifically for echo reports remain limited, and earlier works have not been developed using generic schemas for information extraction^11–13^. Specifically, these studies often focus on a relatively narrow component, such as the comparison statement of reports^13^, or use pre-defined important elements that do not capture the full report content^11,12^. The scarcity of densely annotated reports likely led to reliance on narrow extraction schemas, limiting information scope and hindering the development of comprehensive metrics for assessing factual correctness in clinical echo reports.

RadGraph and RadGraph XL are knowledge graph models addressing similar challenges in radiology report analysis^3,6^. However, despite robust out-of-distribution performance, their direct application to echo reports has significant limitations^3,6,14–16^. Unlike chest X-rays, echo reports frequently contain detailed quantitative measurements that are critical for clinical decision-making, and RadGraph rewards lack the sensitivity to accurately detect and differentiate differences in quantitative measurements^16^. This highlighted the need for a dedicated model tailored specifically to the unique demands of echo reports.

In this context, we plan to develop EchoGraph with a refined clinical entity and relation extraction schema and reward function specifically tailored to echo reports. We hypothesize that the EchoGraph model, along with the new reward, will achieve comparable performance to RadGraph in detecting shared entities and relations while demonstrating enhanced sensitivity in accurately capturing quantitative measurements. This dedicated approach aims to address the unique requirements of unstructured echo report data, ensuring precise evaluation of clinically relevant metrics.

## Results

### Dataset

The Mayo dataset comprises 600 patients with a mean age of 64.9 ± 15.8 years, predominantly white (91.8%), and 54.1% male. A majority of the studies (83.8%) were TTE, followed by transesophageal echocardiography (7.7%). Baseline comorbidities are detailed in **Supplementary Table 1**. The MIMIC-EchoNotes dataset also had 83.3% TTE studies, but demographic and comorbidity information was unavailable for the anonymized data^17^. The Mayo Clinic Health System dataset contains 150 consecutive adult studies, with the predominant modality being TTE (88.7%), followed by stress echocardiography (6.7%) and TEE (4.0%). Among the 150 patients, the mean age was 70.1 ± 12.8, 83 (55.3%) of patients were male, and 147 (98.0%) were white.

### Dataset Annotation

The Mayo dataset has a total of 48,256 entities and 29,731 relations across all 600 reports, and the MIMIC-EchoNotes has a total of 3,672 entities and 2,360 relations across the 60 reports. The most predominant entity category was ‘Anatomy’ (30.3%) and ‘Observation-definitely present’ (39.5%) for the Mayo and EchoNotes datasets, respectively. The frequency of ‘Measurement’ is lower in EchoNotes compared to the reports at Mayo (16.8% vs. 29.3%). Relatively low frequencies of ‘Observation-definitely absent’, ‘Observation-uncertain,’ ‘Located at,’ and ‘Suggestive of’ were observed. Notably, there were no ‘OBS-UC’ examples in our Mayo test set, as determined by our patient-level random split procedure. Details are summarized in **Table 1**.

**Table 1:**
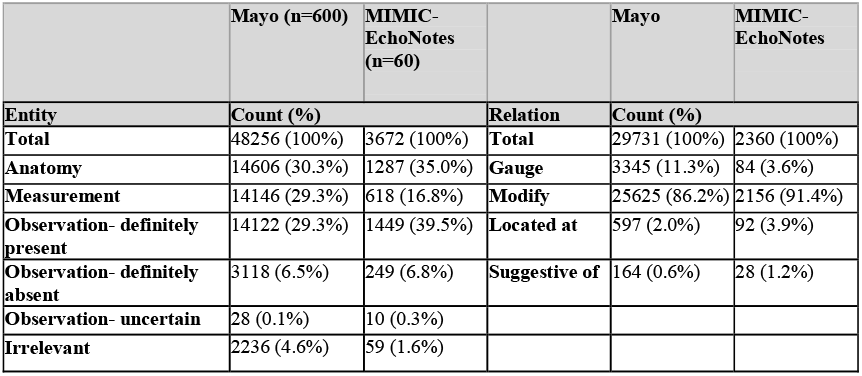
Annotation Counts.

### Schema Coverage

Our information extraction schema resulted in a high percentage of schema coverage. In the Mayo data set, the percentage of annotated tokens (among the whole report tokens) was 79.8 ± 3.9%, and 73.6 ± 7.1% in the MIMIC-EchoNotes dataset, as summarized in **Table 2**.

**Table 2.**
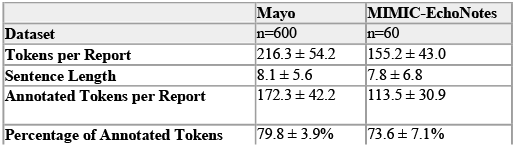
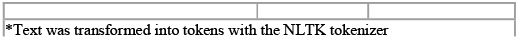
Schema Coverage.

### Interobserver Agreement

The initial annotation yielded an average agreement of 0.82 for entities and 0.60 for relations. After review and consensus among the three annotators, the average agreement improved to 0.91 for entities and 0.71 for relations.

### Model Performance

The EchoGraph model demonstrated robust performance on the Mayo test set for both entity and relation extraction tasks, while also showing generalizability when evaluated on the external MIMIC-EchoNotes dataset (**Table 3**). When examined by individual entities and relations, EchoGraph demonstrated the best performance in identifying Measurement and Observation: Definitely Present on the Mayo dataset. However, its performance on Measurement and Observation: Definitely Absent decreased when evaluated on the MIMIC-EchoNotes dataset. In terms of relations, our model performed well on more common categories (Gauge, Modify, and Located at) while showing lower performance on the “Suggestive of” category. A more significant performance drop of “Located at” and failing in predicting “Suggestive of” were observed when evaluating on the MIMIC-EchoNotes dataset (**Table 4**).

**Table 3.**
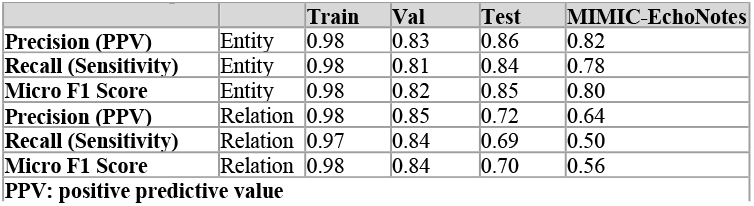
EchoGraph’s Performance.

**Table 4.**
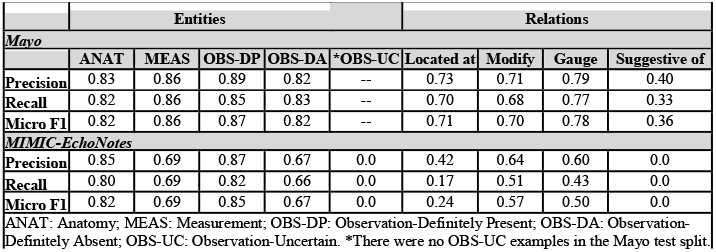
Performance by entities and relations.

### Error Analysis

Error analysis revealed that span mismatch was the predominant error type across all categories of entities (54.9%) and relations (98.2%) on the Mayo dataset (**Supplementary Table 2**). On the MIMIC-EchoNotes dataset, missing entities constituted the primary error type (47.7%), followed by span mismatch (36.1%), while span mismatch remained the dominant error in relation detection (92.8%) (**Supplementary Table 3**).

### EchoGraph F1’s Performance on Corrupted Synthetic Echo Reports

Both EchoGraph F1 and RadGraph F1 achieved perfect scores (1.0) when comparing unaltered reports. EchoGraph F1 demonstrated sensitivity to synthetic reports with 25%, 50%, and 100% corrupted modifiers or numbers (50% and 60% drop on completely corrupted reports, respectively). In contrast, RadGraph F1 showed minimal sensitivity to the corruption in directional terms and measurement numbers, with only modest score reductions (6% and 12% drop, respectively) on completely corrupted reports (**Figure 1**).

**Figure 1.**
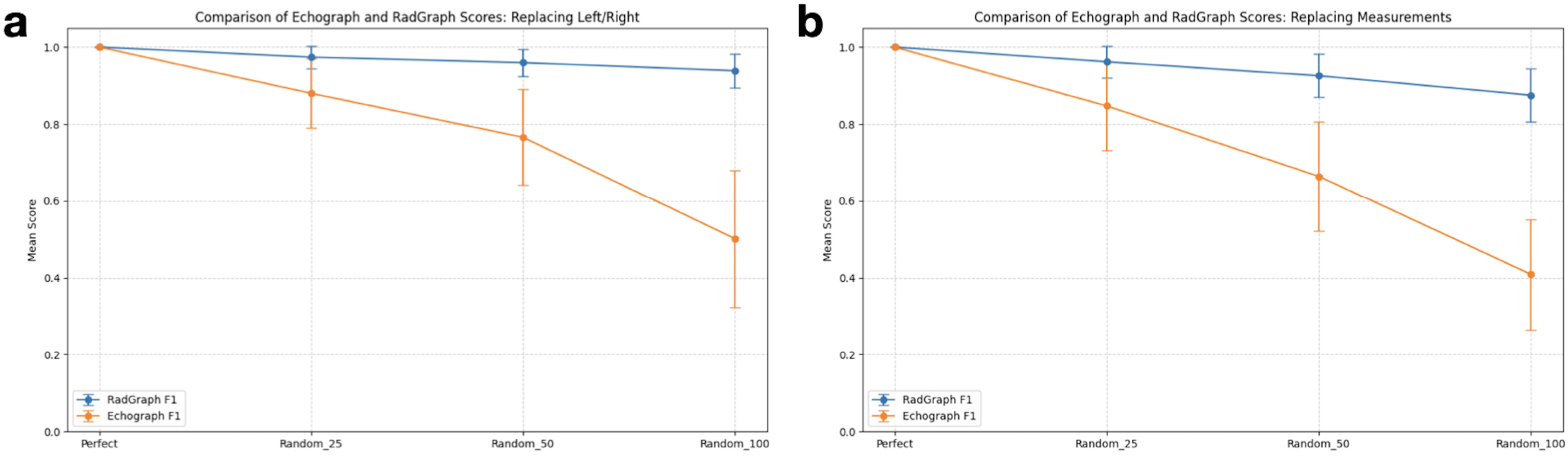
Both RadGraph F1 and EchoGraph F1 achieved perfect scores (1.0) on unaltered reports. **Panel a**: Performance on reports with randomly replaced left and right modifiers. EchoGraph F1 showed high sensitivity to corrupted modifiers (50% drop) while RadGraph F1 maintained near-perfect scores with minimal sensitivity (6% drop). **Panel b**: Performance on reports with randomly replaced measurements. EchoGraph F1 demonstrated significant sensitivity to number corruption (60% drop) while RadGraph F1 showed only modest reduction (12% drop).

### Human Expert Error Annotation Study

The two human experts identified an average of 96% of errors in each report. The two human experts missed an average of 0.35 ± 0.76 errors per report, with an inter-annotator difference of 0.16 ± 1.06 errors. The difference in error sensitivity between EchoGraph F1 and RadGraph F1 was substantial across all metrics. EchoGraph F1 showed 2.8-fold greater slope magnitude (−0.817 vs −0.291), 39% higher variance explained (R^2^ = 0.803 vs 0.578), and stronger correlation (ρ = −0.836 vs −0.715) when assessed against error ratios (detailed in **Table 5**). A representative example is provided in **Supplementary Table 5** to demonstrate the performance of EchoGraph F1 versus RadGraph F1 scores.

**Table 5.**
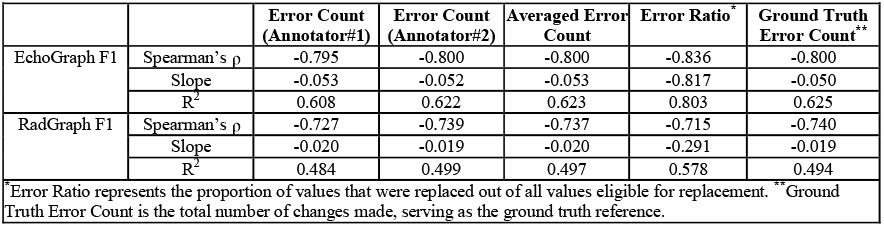
Error sensitivity comparison between EchoGraph F1 and RadGraph F1 scores.

**Table 5.**
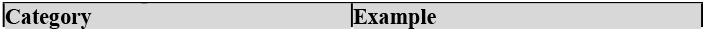
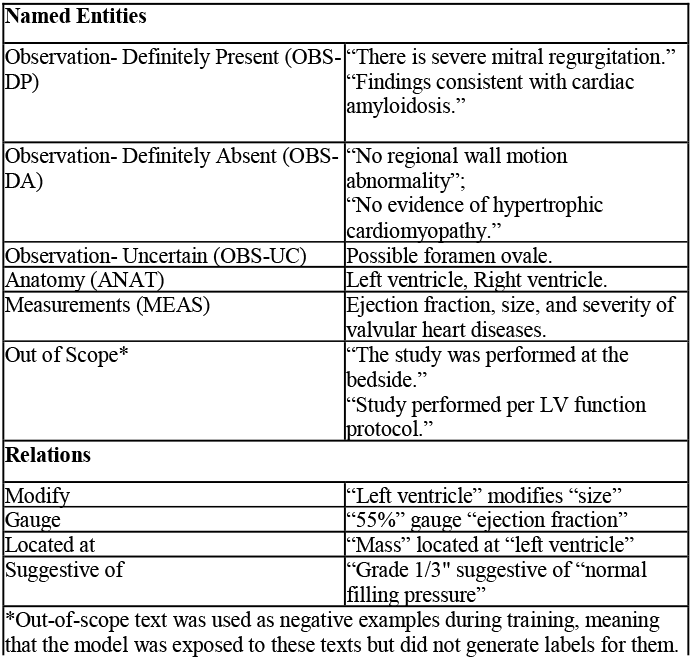
Examples of Entities and Relations.

## Discussion

The contributions of this work are: (1) developing EchoGraph, a BERT-based model with a specialized schema for extracting entities and relations (particularly measurements) from echo reports, and (2) implementing a novel F1-style evaluation metric that prioritizes clinically significant entities and relations for echo reports and demonstrated its strong correlation with human expert preference. EchoGraph overcomes limitations of previous approaches by providing a domain-specific automatic assessment tool for echo report quality.

### Pros and Cons of Expanded Schema

Achieving micro F1 scores of 0.85 and 0.70 in entity and relation prediction tasks, EchoGraph’s performance is close but slightly lower than RadGraph and RadGraph-XL’s (**Tables 3 and 4**)^3,6^. We attribute these results primarily to our expanded extraction schema and the use of phrase-level rather than word-level entity spans. The addition of the measurement category potentially confused the model, as these elements were previously classified under the observation category. The use of phrase-level entity spans was also identified as a common source of disagreement in our annotation quality assessment. For instance, the phrase “calculated 2D-linear” (which modifies ejection fraction) could be annotated as either a single entity or as two separate measurement entities. It is also anticipated that annotations on longer reports are more subject to inter-observer variability^3,6^. Similarly, in our error analysis, we found that a majority of the errors stemmed from the entity span (**Supplementary Table 2**), despite correct entity type predictions.

Overall, EchoGraph demonstrated generalizable performance on the MIMIC dataset, despite dropped performance in low-frequency categories (**Tables 3 and 4**). These low-frequency categories, including “OBS-UC” entity, “Located at” and “Suggestive of” relations, were also found to be low-frequency on reports from other modalities^6^. We observed span-mismatch as the most common error type in relation detection (92.8%), indicating that phrase-level entity spans may be sensitive to slight terminology differences between institutions. Notably, the shorter report and sentence lengths in the MIMIC-EchoNotes dataset pose challenges for relation annotation and detection, as denser annotations are required in limited space.

### An automatic factual correctness metric for AI-generated echo reports

Established natural language processing metrics such as BLEU, ROUGE, and METEOR^20–22^ were not originally designed for evaluating medical texts^16,23^. While RadGraph has provided a strong foundation for the evaluation of radiology reports, its scoring mechanism has limitations when applied to domains like echo reports^16^, where numerical precision and relational semantics are critical.

RadGraph’s uniform treatment of all entity types—including anatomical terms, modifiers, and numeric values—fails to account for the disproportionate clinical significance of certain measurements in echo. Furthermore, RadGraph’s independent treatment of relations disregards the semantic dependency structure between entities, enabling spurious matches when core numeric content is inaccurate (**Figure 1**)^16^. By incorporating a category specifically tailored for measurements and corresponding relations in echo reports, EchoGraph more accurately reflects the semantic coherence and numeric precision essential for clinically meaningful echo reporting. This enhanced clinical relevance is reflected in the substantially stronger error sensitivity of EchoGraph F1 compared to RadGraph F1 when validated against human error annotations in a contemporary dataset (**Table 5**).

Similar to its predecessor models^3,6^, EchoGraph can be leveraged for similar tasks, including automatic annotation of large-scale unstructured echo report text, to evaluate and improve the quality of AI-generated echo reports^3,6,16^, as well as enhancing vision-language models in identifying fine-grained pairs between images and corresponding text^24^.

The EchoGraph study is limited by its retrospective nature. The development and external validation datasets were both from tertiary referral medical centers and may not be representative of the general population or reporting styles at different institutions. The development dataset was from 2017 and may not reflect contemporary echocardiography reporting language. Due to the need for dense annotations, obtaining large-scale datasets at the patient level is challenging, which substantially limits the representativeness of patient variability. All standard abbreviations were expanded for the consistency of annotation, which may hinder the model’s ability to identify abbreviations. Additionally, both development and validation datasets were in English, which limits EchoGraph’s application to non-English reports. EchoGraph is also subject to limitations such as dependence on text order and exact text match. For example, “no significant valvular diseases” is equivalent to a ground truth of “trivial mitral and tricuspid regurgitation,” yet this could be rated low. Such issues could be addressed by language models that understand the clinical meaning of these equivalent statements with specific fine-tuning^7^. As a BERT-based model, EchoGraph is smaller and faster than large language models^14,15,25^, requiring fewer computational resources as a cost-effective solution for report quality assessments. Although phrase-level entity annotation introduced variation that impacted the model’s performance on external datasets, this approach was chosen for its feasibility, considering the relatively long context of the reports.

In conclusion, EchoGraph advances automatic echo report evaluation through its specialized entity-relation extraction schema and clinically weighted rewards that prioritize measurements. This approach provides an effective and scalable solution for evaluating and enhancing the development of language model-based applications in echo.

## Methods

### Mayo Clinic Echo Reports

Adult (> 18 years old) echo studies performed from 1/1/2017 to 12/31/2017 at the Mayo Clinic Enterprise were retrieved, and 600 reports from non-duplicated patients were randomly selected. The Findings section was used to ensure all relevant clinical observations were considered. This section contains detailed clinically significant statements, including key quantitative parameters such as left ventricular ejection fraction, global longitudinal strain, left atrial volume index, right ventricular systolic pressure, ascending aortic dimensions, and transvalvular gradients. Subheadings were removed from the text. Following this, the dataset was split into the train, validation, and test sets in a 7:2:1 ratio. A validation set (n=1,000) from Mayo Arizona was randomly selected for the data corruption study; the characteristics of this set were described in our previous work^16^. To investigate the correlation between EchoGraph and human expert preferences in contemporary echocardiography reports, we collected 150 consecutive adult patients from the Mayo Clinic Health System in 2023.

### MIMIC-III ECHO-NOTE2NUM Reports

(v.1.0.0, referred to as MIMIC-EchoNotes below)^17^: This publicly available dataset contains 43,472 valid free-text echo reports from the intensive care unit at the Beth Israel Deaconess Medical Center between 2001 and 2012. Similarly, the Findings section of a random subset (n=60) was annotated and used for external validation. Common abbreviations in the text were expanded to their full forms.

### Dense Named Entity and Relation Annotation

The annotation schema covers both named entities and relations, as detailed in **Table 5** with corresponding examples. In brief, named entities include “Observation-Definitely Present” (OBS-DP), “Observation-Definitely Absent” (OBS-DA), “Observation-Uncertain” (OBS-UC), “Anatomy” (ANAT), and “Measurements” (MEAS), while relations encompass “Modify,” “Gauge,” “Located at,” and “Suggestive of.” Relations were only considered within sentences.

While largely adopting the extraction schema of RadGraph^3^, we specifically designed “MEAS” and “Gauge” to reflect the requirements for assessment and grading in echo reports (**Figure 2**).

**Figure 2.**
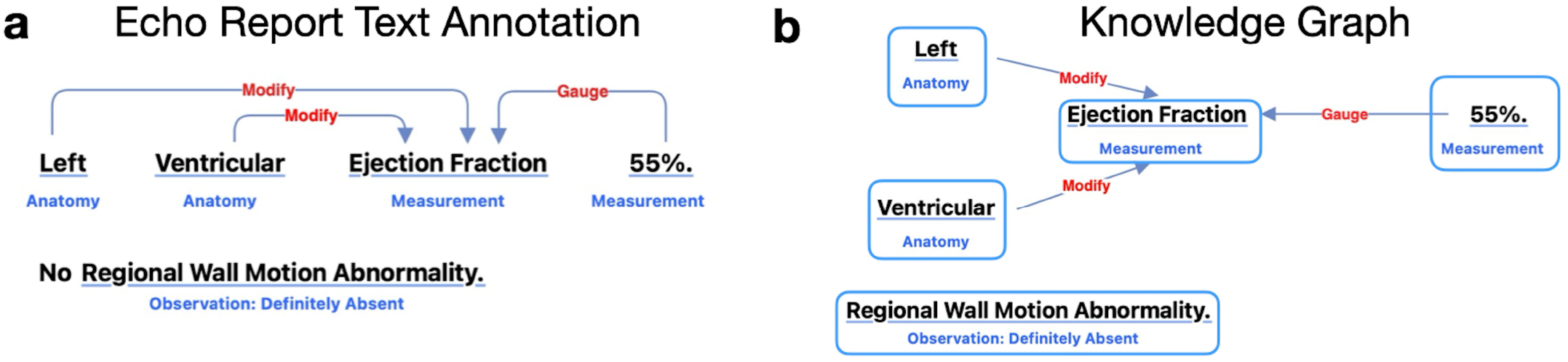
A depiction of the text annotation of an echo report and the corresponding knowledge graph. **Panel a** illustrates the annotation of two sentences: “left ventricular ejection fraction 55%” and “no regional wall motion abnormality.” Here, “left” and “ventricular” are annotated as anatomy entities, while “ejection fraction” and “55%” are annotated as measurement entities. “No regional wall motion abnormality” is annotated as Observation: Definitely Absent. The relations between entities are marked as “Modify” and “Gauge.” **Panel b** shows the corresponding knowledge graph for this annotation. Notably, “regional wall motion abnormality” is isolated from other entities as no relations are annotated between the two sentences.

To accommodate the extended length of echo reports while conforming to our schema requirements, our annotation strategy targeted phrase-level entities such as “left ventricle,” “left atrium,” and “regional wall motion abnormality” instead of individual words, simplifying the annotation process. Statements in the report that were unrelated to diagnosis, such as quality control notes (e.g., “study performed at the bedside”), were classified as out-of-scope irrelevant and not used for training. The labels of out-of-scope irrelevant were then removed to use the corresponding text as negative examples for model training. An open-source annotation platform, Doccano, was used for annotation (https://github.com/doccano/doccano). The raw annotations were used to train the model without further processing.

### Annotating Consistency and Interobserver Agreement

As part of the annotation quality control process, the three annotators (FG, AKM, MTA) all independently annotated a small sample (n=10) from the Mayo dataset to evaluate the interobserver agreement, and the consistency of the annotations was reviewed by senior cardiologists (JFM and CJC) and revised according to the group consensus. The agreement, defined as an exact match of annotations for entities (e.g., 3-element list of [“start word index”, “end word index”, “label”]) and relations (e.g., 5-element list of [“from start word index”, “from end word index”, “to start word index”, “to end word index”, “type”]) were averaged over all annotated samples.

### Model Training

We fine-tuned a pre-trained Bidirectional Encoder Representations from Transformers (BERT)-based model (sciBERT)^18^ using the DYGIE++ framework for joint entity and relation extraction^3,6^. The framework was selected due to its effectiveness in similar studies^3,6^. Specifically, the model was trained for 50 epochs using a learning rate of 5e-5, weight decay of 0.01, and batch size of 1 on a 24GB NVIDIA RTX A5000 GPU.

### Model Performance Evaluation

We adopted the evaluation approach in the RadGraph study^3^ to report the micro F1 for named entity recognition and relation extraction, concerning the imbalanced distribution between classes. For entity prediction, a correct prediction is defined as an exact match of the span boundaries and entity type between the model’s prediction and the annotation (e.g., [1, 2, ANAT] matches [1, 2, ANAT]). Similarly, for relation extraction, a correct prediction requires an exact match of the span boundaries of the entity pair and the relation type (e.g., [1, 2, 3, 4, Modify] matches [1, 2, 3, 4, Modify]).

### EchoGraph F1 score

An F1 score can be derived from the predictions of the EchoGraph model to automatically assess echo report quality against reference reports. We introduced a new F1-style reward, “weighted entity-incoming relations reward”, for EchoGraph’s reward calculation. This reward adopts a node-centric subgraph approach by defining “central entities”, which are entities without outgoing relations, and gives more weight to other entities with direct relations to the main entities (**Figure 3**).

**Figure 3.**
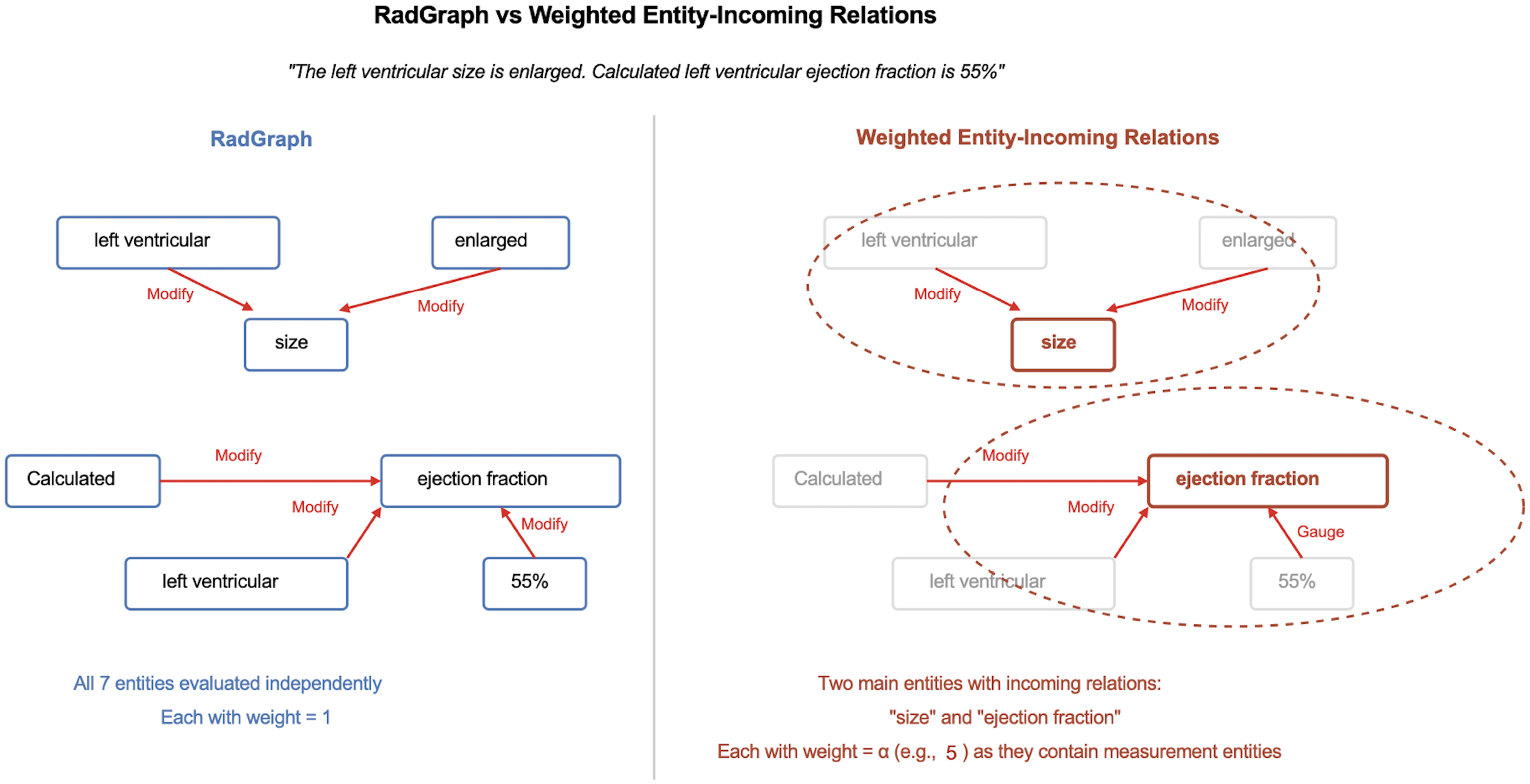
Illustration comparing RadGraph’s independent entity evaluation versus EchoGraph’s weighted entity-relation reward approach. Using a sample sentence describing left ventricular size and ejection fraction measurements, RadGraph evaluates detected entities independently without additional weighting. In contrast, EchoGraph identifies main entities (typically measurements) and assigns a desired weight (here = 5) to their incoming relations during reward calculation.

To emphasize the critical role of numeric measurements, these enriched semantic units are assigned higher weights if the central entity itself or any entity with incoming relations has a numeric measurement (label: MEAS).

Formally, each enriched entity *e* is assigned a weight *w*(*e*):

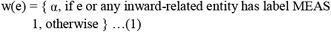

where *α* > 1(5 was used in our experiments), thereby prioritizing the accuracy of numeric measurements. Precision and recall are computed using these weighted counts:

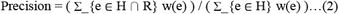

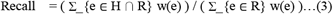

The weighted F1-score is then:

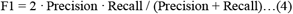

### The Impact of Data Corruption on Evaluation Metrics in Echo Reports

We examined how automated evaluation metrics respond to corrupted data in echo reports. Using a dataset of 1,000 Mayo Clinic echo reports from Arizona^16^, we systematically replaced measurement values and directional terms (“left”/”right”) at varying degrees (25%, 50%, and 100%). We then analyzed how these manipulations affected Echograph F1 and RadGraph F1 scores when comparing the altered reports against their original versions.

### Error Analysis

An error analysis was conducted to identify whether the source of the error originated from the entity span or the prediction label, or was completely missing. The definition of mismatch categories is provided in **Supplementary Table 4**.

### Human Expert Error Annotation Study

Similar to the data corruption study, the Findings section of each report was randomly modified by altering 0%, 50%, 75%, or 100% of key information, including measurement values, normal versus abnormal classifications, and severity modifiers (e.g., mild replaced by severe). Two cardiology experts independently annotated the number of errors in each report compared to the original. The relationship between error counts and both EchoGraph F1 and RadGraph F1 scores was assessed using Spearman’s correlation, linear regression slope (score change per error), and R^2^ (variance explained by errors).

### Ethical review and approval

#### All the studies have been performed in accordance with the Declaration of Helsinki

The use of the Mayo Clinic dataset was approved by the institutional review board (protocol#22-010944); only patients providing informed consent for minimal-risk retrospective studies were included, thus the requirement for additional informed consent was waived. The MIMIC-EchoNote dataset is publicly available and was granted by the Institutional Review Boards of Beth Israel Deaconess Medical Center and the Massachusetts Institute of Technology. Patient consent was waived, as the project had no bearing on clinical care, and all protected health information was de-identified^17^.

## Data Availability

We released EchoGraph-annotated MIMIC-EchoNote data to facilitate future research (https://physionet.org/PrhchnmblxOSQ5AgK7czF6Udum8eorEGQBQZxHWxxysJfEWE4QlQ2yLC3wlX9qpi/). The Mayo Clinic data set is not publicly available due to patient privacy considerations, however, it can be obtained from the corresponding author upon reasonable request.

## Code Availability

The EchoGraph model is released at: https://github.com/chiehjuchao/echograph

## Abbreviations

AI: Artificial Intelligence
DL: Deep Learning
Echo: Echocardiography
FT: Fine-Tuning
GPT: Generative Pre-trained Transf
Seq2seq: Sequence-to-sequence
TTE: Transthoracic Echocardiography
TEE: Transesophageal Echocardiography
LLM: Large Language Model

## Acknowledgments

Chieh-Ju Chao, MD, is supported by research funding from the CV Prospective award from the Mayo Clinic Department of Cardiovascular Medicine and the AI/ML Enablement award from the Center for Digital Health at the Mayo Clinic. The other authors declare no relevant funding for this work.

## Author Contributions

C.J.C.: Conceptualization, methodology, software, validation, data curation, formal analysis, writing – original draft, visualization.

J.B.D.: Methodology, software.

M.A.: Methodology, software.

I.B.: Methodology, resources, writing – review and editing.

J.M.F.: Data curation.

F.G.: Data curation.

A.K.M.: Data curation.

M.T.A.: Data curation.

Y.C.W: Data curation.

R.A.: Resources, writing – review and editing.

G.C.K.: Resources, writing – review and editing.

J.K.O.: Resources, writing – review and editing.

B.J.E.: Resources, writing – review and editing.

F.F.L.: Methodology, resources, writing – review and editing, supervision.

E.A.: Conceptualization, methodology, resources, writing – review and editing.

C.P.L.: Conceptualization, resources, writing – review and editing, supervision. All authors have read and approved the manuscript.

