## Supplemental Table 1 for "EchoGraph System for Automated Quality Assessment of Echocardiography Reports"

### Supplementary Materials

| Supplementary Table 1. Patient Demographics | | |
|---|---|---|
| | Mayo | MIMIC-EchoNote |
| | n=600 | n=60 |
| **Age** | 64.9 ± 15.8 | – |
| **Race** | | |
| ***White*** | 551 (91.8%) | – |
| ***Black*** | 19 (3.2%) | – |
| ***Other*** | 20 (3.3%) | – |
| ***Asian*** | 6 (1.0%) | – |
| ***Native American*** | 3 (0.5%) | – |
| ***Pacific Islander*** | 1 (0.2%) | – |
| **Sex** | | |
| ***Male*** | 324 (54.1%) | – |
| ***Female*** | 275 (45.9%) | – |
| **HTN** | 82 (13.7%) | – |
| **DM** | 100 (16.7%) | – |
| **CAD** | 49 (8.2%) | – |
| **CKD** | 104 (17.3%) | – |
| **Stroke** | 24 (4.0%) | – |
| **Echo Study Type** | | |
| ***Adult TTE*** | 502 (83.8%) | 50 (83.3%) |
| ***Adult TEE*** | 46 (7.7%) | 10 (16.7%) |
| ***Stress Echo*** | 51 (8.5%) | – |

**Supplementary Table 2. Error Analysis on the Mayo Dataset**

| Issue Type | ANAT | MEAS | OBS-DA | OBS-DP | OBS-UC | Total |
|---|---|---|---|---|---|---|
| **Entity** | | | | | | |
| Missing | 38 (13.5%) | 38 (18.4%) | 5 (8.8%) | 49 (21.9%) | – | 130(16.9%) |
| Span and type mismatch | 115 (40.8%) | 13 (6.3%) | 17 (29.8%) | 47 (21.0%) | – | 192(24.9%) |
| Span mismatch | 128 (45.4%) | 150 (72.5%) | 34 (59.6%) | 111 (49.6%) | – | 423(54.9%) |
| Type mismatch | 1 (0.4%) | 6 (2.9%) | 1 (1.8%) | 17 (7.6%) | – | 25(3.2%) |
| **Relation** | | | | | | |
| | **Gauge** | **Located at** | **Modify** | **Suggestive of** | | **Total** |
| Missing | 6 (7.6%) | 4 (26.7%) | 0 (0.0%) | 3 (25.0%) | – | 13 (1.4%) |
| Span and type mismatch | 0 (0.0%) | 1 (6.7%) | 0 (0.0%) | 1 (8.3%) | – | 2 (0.2%) |
| Span mismatch | 73 (92.4%) | 10 (66.7%) | 851 (99.8%) | 8 (66.7%) | – | 942 (98.2%) |
| Type mismatch | 0 (0.0%) | 0 (0.0%) | 2 (0.2%) | 0 (0.0%) | – | 2 (0.2%) |

ANAT: Anatomy; MEAS: Measurement; OBS-DP: Observation-Definitely Present; OBS-DA: Observation-Definitely Absent; OBS-UC: Observation-Uncertain.

| Supplementary Table 3. Error Analysis on EchoNote-MIMIC Dataset | | | | | | |
|---|---|---|---|---|---|---|
| **Issue Type** | **ANAT** | **MEAS** | **OBS-DA** | **OBS-DP** | **OBS-UC** | **Total** |
| **Entity** | | | | | | |
| Missing | 157 (57.7%) | 91 (44.0%) | 17 (18.5%) | 138 (50.2%) | 5 (50.0%) | 408 (47.7%) |
| Span and type mismatch | 37 (13.6%) | 6 (2.9%) | 38 (41.3%) | 11 (4.0%) | 0 (0.0%) | 92 (10.7%) |
| Span mismatch | 78 (28.7%) | 94 (45.4%) | 32 (34.8%) | 105 (38.2%) | 0 (0.0%) | 309 (36.1%) |
| Type mismatch | 0 (0.0%) | 16 (7.7%) | 5 (5.4%) | 21 (7.6%) | 5 (50.0%) | 47 (5.5%) |
| **Relation** | | | | | | |
| | **Gauge** | **Located at** | **Modify** | **Suggestive of** | | **Total** |
| Missing | 6 (12.8%) | 36 (47.4%) | 0 (0.0%) | 14 (50.0%) | – | 56 (4.7%) |
| Span and type mismatch | 3 (6.4%) | 5 (6.6%) | 0 (0.0%) | 14 (50.0%) | – | 22 (1.9%) |
| Span mismatch | 37 (78.7%) | 33 (43.4%) | 1033 (99.5%) | 0 (0.0%) | – | 1103 (92.8%) |
| Type mismatch | 1 (2.1%) | 2 (2.6%) | 5 (0.5%) | 0 (0.0%) | – | 8 (0.7%) |

ANAT: Anatomy; MEAS: Measurement; OBS-DP: Observation-Definitely Present; OBS-DA: Observation-Definitely Absent; OBS-UC: Observation-Uncertain.

**Supplementary Table 4. Definition of Mismatch Categories**

| **Mismatch Categories** | **Definition (Entity)** | **Definition (Relations)** |
|---|---|---|
| Span | Incorrect starting OR ending word index | Incorrect starting OR ending entity span |
| Type | Incorrect label | Incorrect label |
| Span and Type | Incorrect starting OR ending word index, AND incorrect label | Incorrect starting OR ending entity span, AND incorrect label |
| Missing | Incorrect starting AND ending word index AND incorrect label | Incorrect starting AND ending entity span AND incorrect label |

| Supplementary Table 5. Representative Example of Synthetic Error Report | | | |
|---|---|---|---|
| **Original Report** | **Modified Report*** | **EchoGraph F1 score** | **RadGraph F1 score** |
| - Normal left ventricular chamber size.<br>- Normal left ventricular systolic function.<br>- Calculated M-mode left ventricular ejection fraction 65%.<br>- Normal right ventricular chamber size.<br>- Normal right ventricular systolic function.<br>- Normal cardiac valves.<br>- Positive for atrial level shunt by color flow imaging. - Small atrial septal defect.<br>- Left-to-right shunt at atrial level.<br>- No shunt at ventricular level.<br>- Normal scan of the aorta.<br>- No pericardial effusion. | - Abnormal left ventricular chamber size.<br>- Abnormal left ventricular systolic function.<br>- Calculated M-mode left ventricular ejection fraction 47%.<br>- Abnormal right ventricular chamber size.<br>- Abnormal right ventricular systolic function.<br>- Normal cardiac valves.<br>- Positive for atrial level shunt by color flow imaging.<br>- Small atrial septal defect.<br>- Left-to-right shunt at atrial level.<br>- No shunt at ventricular level.<br>- Normal scan of the aorta.<br>- No pericardial effusion. | 0.21 | 0.67 |
| *Errors in red font. | | | |
